# Effectiveness of BNT162b2 mRNA vaccine and ChAdOx1 adenovirus vector vaccine on mortality following COVID-19

**DOI:** 10.1101/2021.05.14.21257218

**Authors:** Jamie Lopez Bernal, Nick Andrews, Charlotte Gower, Julia Stowe, Elise Tessier, Ruth Simmons, Mary Ramsay

## Abstract

We estimated risk of death in vaccinated compared to unvaccinated COVID-19 cases. Cases vaccinated with 1 dose of BNT162b2 had 44% reduced risk of death, 55% with 1 dose of ChAdOx1, and 69% with 2 doses of BNT162b2. This is on top of the protection provided against becoming a case.

## Introduction

Real world effectiveness data has begun to emerge for COVID-19 vaccines, showing high levels of protection against both symptomatic and asymptomatic infection, supporting the findings of the phase III clinical trials.(1–6) Nevertheless, evidence on effectiveness against mortality, the most severe outcome, is currently limited and has not yet been reported for most vaccines.(2)

The UK was the first country to implement a COVID-19 vaccination programme, starting with the Pfizer-BioNTech BNT162b2 mRNA vaccine in December and followed soon afterwards with the Oxford-Astrazeneca ChAdOx1 adenovirus vector vaccine. Vaccination was offered to older adults, care home residents and health and social care workers first, delivery was subsequently rolled out to increasingly younger age cohorts and to clinical risk groups. A policy decision was made early on in the programme to use an extended dosing interval of 12 weeks, in order to maximise the number of vulnerable people offered the first dose.

Early data suggested that a single dose of BNT162b2 was 80-85% effective at preventing mortality in individuals aged over 80 years.(2) However, effectiveness of ChAdOx1 against mortality has not yet been reported.

## Methods

This study estimates the risk of death among confirmed cases of COVID-19 according to their vaccination status.

In England a community COVID-19 testing programme is available to individuals reporting symptoms (high temperature, new continuous cough, loss or change in sense of smell or taste), care home residents and staff, and individuals taking part in local or national mass asymptomatic testing. We linked all new symptomatic PCR positive cases to mortality data from NHS records.(7) Testing and deaths data between 8^th^ December (when the vaccination programme started) and 17^th^ April were initially extracted. This was subsequently restricted to deaths to April 6^th^ as lags in deaths registrations could affect the most recent days.

Survival analysis was conducted to estimate the hazard ratios for death within 28 days of a positive SARS-CoV-2 PCR test by vaccination status: unvaccinated, vaccinated with one dose (where the first dose was received at least 21 days before the test date), vaccinated with 2 doses (where the second dose was received at least 7 days before). We also estimated effects among those who received first doses 0-20 days prior to testing positive (earlier than an effect would be anticipated) as a control analysis. Adjustments were made for sex, clinical risk factors for severe disease (including immunosuppressive disease or therapy, severe respiratory disease, rare diseases and inborn errors of metabolism that significantly increase the risk of infections, pregnancy with significant congenital heart disease),(8) age in 5 year bands, and whether they had been identified as a care home resident. Estimates were stratified by age group (70-79 and 80+ years) and whether they were a care home resident.

## Results

48,096 cases aged 70 years and above were included in the analysis, of which 79.1% were unvaccinated; 12.7% had been vaccinated with BNT162b2 (3,910 received their first dose within 20 days of their test date, 2,007 received their first dose >=21 days before their test date and 191 received their 2^nd^ dose >=7 days before their test date); and 8.2% had been vaccinated with ChAdOx1 (2,686 received their first dose within 20 days of their test date, 1,258 received their first dose >=21 days before their test date and 6 received their 2^nd^ dose >=7 days before their test date).

Kaplan Meier curves showing the proportion who died by days after the positive test for the first dose and unvaccinated are shown in figure 1. Among 80+ year olds with at least 28 days of follow-up data 16.1% (1462/9105) of unvaccinated cases died, 9.2% (99/1,072) of cases vaccinated with 1 dose of BNT162b2 died, 11.3% (33/293) of cases vaccinated with 1 dose of ChAdOx1, and 4.7% (6/128) of cases vaccinated with 2 doses of BNT162b2 died. Among 70-79 year olds with at least 28 days of follow-up data 4.0% (1,147/28,875) of unvaccinated cases died, 2.7% (15/549) of cases vaccinated with 1 dose of BNT162b2 died, 2.1% (10/484) of cases vaccinated with 1 dose of ChAdOx1, and 0% (0/7) of cases vaccinated with 2 doses of BNT162b2 died. There were no individuals with 28 days of follow-up after 2 doses of ChAdOx1.

**Figure 1:**
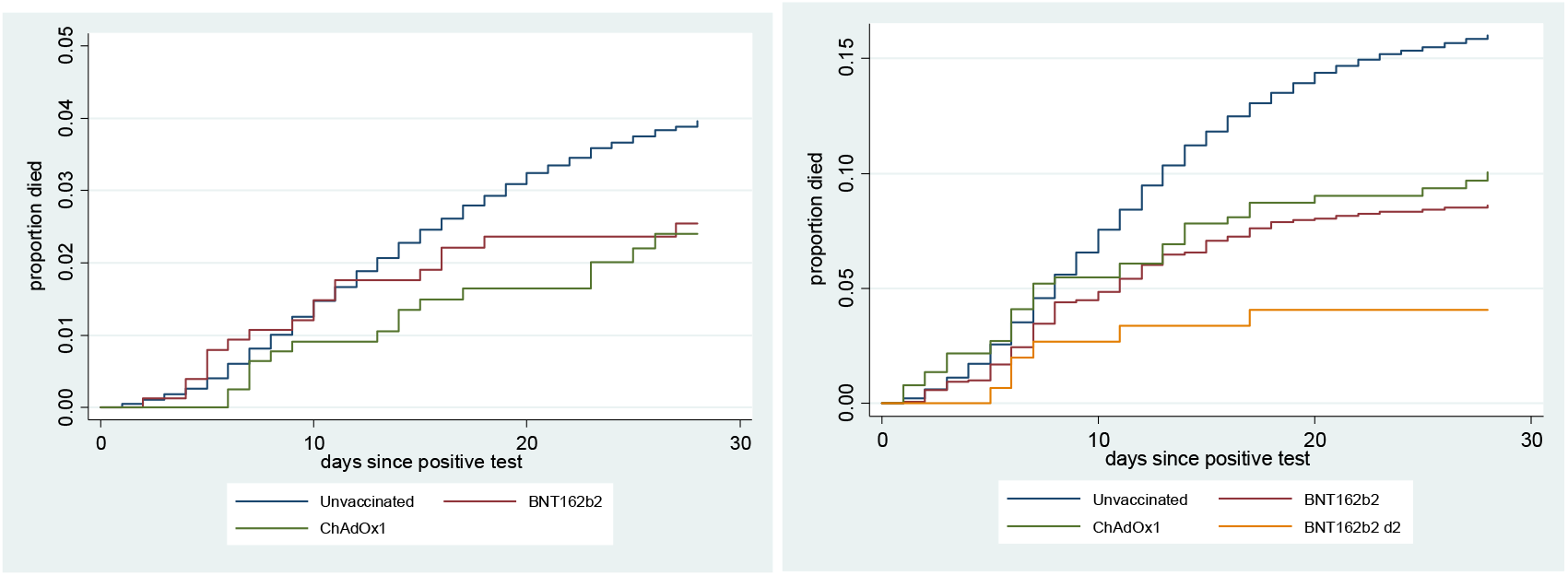
Proportion died by vaccination status (a) 70-79 year olds (b) 80+ year olds

In the fully adjusted model, hazard ratios compared to unvaccinated cases were 0.56 (95%CI 0.47-0.68) for 1 dose of BNT162b2, 0.45 (95%CI 0.34-0.59) for 1 dose of ChAdOx1 and 0.31 (95%CI 0.14-0.69) for 2 doses of BNT162b2. This indicates an additional 44% protection against death after the first dose of BNT162b2, 55% after the first dose of ChAdOx1, and 69% after the second dose of BNT162b2, on top of the protection against become a case. There was little difference by age group; the hazard ratios after the first BNT162b2 dose were 0.49 (95% CI 0.31-0.78) and 0.56 (95% CI 0.46-0.69) for age 70-79 and 80+, respectively, whilst for ChAdOx1 they were 0.46 (95%CI 0.28-0.75) and 0.45 (95% CI 0.32-0.63) respectively. Note that in age 80+ care home adjustment was important and led to a lower hazard ratio for ChAdOx1 despite the crude Kaplan-Meier showing lower mortality for BNT162b2.

In the control analysis among those vaccinated with first doses 0-20 days prior to testing positive, the hazard ratio with BNT162b2 was 0.81 (95% 0.73-0.91) and with ChAdOx1 0.99 (95%CI 0.87-1.12).

There were notable differences by care home resident status. Among care home residents, hazard ratios were 0.87 (95%CI 0.60-1.25) for 1 dose of BNT162b2 and 0.37 (95%CI 0.23-0.60) for 1 dose of ChAdOx1 (numbers were too small for estimating 2 dose effects with either vaccine). Among non-care home residents, hazard ratios compared to unvaccinated cases were 0.48 (95%CI 0.38-0.59) for 1 dose of BNT162b2 and 0.52 (95%CI 0.37-0.73) for 1 dose of ChAdOx1.

## Discussion

We found that confirmed cases of COVID-19 who had been vaccinated with either a single dose of BNT162b2 or a single dose of ChAdOx1 had significantly reduced risk of dying compared to unvaccinated cases. The point estimates showed greater protection with ChAdOx1, though confidence intervals overlapped suggesting little difference between the two vaccines. Combined with recent estimates of protection against symptomatic disease in the same age group, these findings suggest that a single dose of either vaccine is approximately 80% effective at preventing mortality, and two doses of BNT162b2 is approximately 97% effective at preventing mortality in older adults.(2, 9)

The phase three clinical trials have been unable to report on efficacy against mortality due to smaller numbers. Our findings with BNT162b2 were similar to real world findings previously reported in the UK and Israel, both in the context of B.1.1.7 variant as the dominant strain.(2, 10) Our study is the first to estimate for the effectiveness of ChAdOx1 against mortality and suggests that a single dose offers similar levels of protection to BNT162b2.

We found that although both vaccines provided additional protection against mortality overall, among care home residents who received a single dose of the Pfizer vaccine, the effect was small and non-significant suggesting that there was no clear evidence additional protection (beyond the protection against becoming a case). Though this could be due to residual confounding and lacking statistical power in this group.

Our study is observational and as such we are unable to exclude unmeasured or residual confounding. For example, healthy vaccinee effects, whereby healthier individuals are more likely to present for vaccination and also more likely to survive, could be an issue. We see some evidence of a possible healthy vaccinee effect with the BNT162b2 because we see a small effect in days 0-20 after the first dose, before a vaccine effect might be anticipated, whereas we don’t see this effect with ChAdOx1. However, we have previously found that vaccine effects occur more rapidly with BNT162b2, so this could represent early vaccine effects.(2) A further limitation is that some of the deaths within 28 days of a positive test may be unrelated to COVID and therefore not preventable by vaccination. All of the individuals included in this analysis had reported symptoms of COVID-19, therefore the proportion of deaths unrelated to COVID is likely to be small, however, this could cause us to underestimate the effects on mortality.

In conclusion, we find strong evidence that both vaccines offer high levels of protection against mortality after a single dose which supports prioritising the first dose in at risk groups in the context of high disease incidence and vaccine supply or delivery constraints. Nevertheless, a second dose of BNT162b2 offers yet further protection against mortality, highlighting the importance of completing the course. We are not yet able to estimate two dose effects with ChAdOx1 due to the later rollout of this vaccine.

## Data Availability

All relevant data is included in the manuscript

